# Effects of dexmedetomidine use on elderly patient’s renal function submitted to laparoscopic surgery: systematic review

**DOI:** 10.1101/2025.05.13.25327563

**Authors:** Yuri Nicolay Kretchetoff, Pedro Emmanuel Alvarenga Americano do Brasil

## Abstract

**Introduction:** Postoperative acute renal failure is a common complication found in patients after laparoscopic surgery. This study aims to systematically review the literature to check the effect of dexmedetomidine on acute renal failure.

**Methods:** A systematic review was carried out, including abstracts published from Jan 2016 to August 2024, where the population of interest was patients aged 60 years or older undergoing laparoscopic surgery. The intervention of interest was dexmedetomidine individually or as an anesthetic adjuvant used in the perioperative period at any dose. The comparison of interest was placebo or any perioperative medication used for the same purpose. Acute renal failure, urinary output, and length of stay (LOS) were the outcomes of interest. The studies of interest were clinical trials. Remote database searches were performed at PubMed, SCOPUS, Web of Science, Embase, and Cochrane, as well as full manuscript bibliographies. The risk of bias was assessed with Cochrane’s RoB2 and results confidence was assessed with GRADE.

**Results:** 13 studies were analyzed. A consistent but not significant nephroprotective effect of dexmedetomidine was observed. A neutral effect was observed in urinary output and LOS, and there is evidence of reporting bias for LOS. The confidence in these findings is either poor or moderate.

**Conclusion:** The use of DEX in the elderly population undergoing laparoscopy surgeries did not reduce the LOS or urinary output, but there is moderate confidence that it has a potential nephroprotective effect.

## Introduction

Laparoscopic surgery in elderly patients has more complications than in younger populations. Studies report overall complication rates ranging from 17.9% to 23.68% in older patients,^[1]^ depending on advanced age, diagnostic criteria, vasopressors, mechanical ventilation, etc. ^[1]^ Minor changes in creatinine and urea after surgery may increase the length of hospital stay (LOS), mortality, the risk of chronic kidney disease (CKD), and the need for hemodialysis.^[2,3]^

The etiology of laparoscopic perioperative AKI is complex and can be triggered by hypoperfusion or neuroendocrine response to surgical stimulus. Frequently it is associated with hypotension and hypoperfusion. It is also related to vasopressor use, although the renal autoregulation mechanism efficiently maintains glomerular filtration.^[2]^

In the inflammatory process, prostaglandins reduce afferent arteriolar resistance, maintaining glomerular blood flow, activating the renin-angiotensin-aldosterone system, and the release of angiotensin II. This results in efferent arteriolar vasoconstriction and maintenance of adequate glomerular pressure in states of renal hypoperfusion. If this state persists or hypovolemia is greater than the capacity to maintain renal perfusion, the autonomic nervous system generates afferent arteriolar vasoconstriction in an attempt to maintain adequate glomerular pressure. This leads to reduced renal blood flow, ischemia, and reduced glomerular filtration.^[2]^ There is also the impact on renal function due to reduced blood flow to the kidneys by mechanical compression of the abdominal vessels due to laparoscopic pneumoperitoneum.^[2]^

Alpha receptor agonists were introduced in anesthetic practice long ago. They modulate the sympathetic and endocrine metabolic response.^[4]^ Dexmedetomidine (DEX) has both peripheral and central action, centrally inhibiting the locus coeruleus and the exocytosis of norepinephrine, leading to hypotension and bradycardia. The stimulation of α2-agonist receptors decreases sympathetic outflow, reducing circulating serum catecholamine levels, leading to a predominance of parasympathetic action and resulting in lower blood pressure. ^[4]^ The most common adverse reactions to the use of DEX are hypotension (30%), hypertension (12%), nausea (11%), bradycardia (9%), and dry mouth (3%). The increase in DEX concentrations progressively decreases heart rate and cardiac output, and there is a potential for asystole, although rare. ^[4]^

DEX has been tested for several purposes, such as sedation in general anesthesia,^[5]^ prolonging the effect of peripheral blocks,^[6]^ postoperative delirium,^[7]^ reducing opioid consumption,^[5]^ reducing nausea and vomiting, and providing good control of hemodynamic response. Clonidine (an alpha-2 blocker) can prevent renal function deterioration in patients undergoing cardiac surgery.^[5]^ More recently, there is evidence of the nephroprotective effect of continuous infusion of DEX during cardiac surgeries.^[6]^ Also, DEX has benefits on overall surgery with general anesthesia in an overall population by reducing hospital mortality, AKI, and intensive care unit (ICU) LOS.^[8]^

This research aims to systematically review the medical literature and trial registries regarding the effect of DEX on AKI, mortality, and LOS when used on elderly populations submitted to laparoscopy.

## Materials and Methods

This systematic review was registered in the International Prospective Register of Systematic Reviews (PROSPERO) under the ID CRD42023456529.

### Eligibility criteria

Population: the population of interest was patients aged 60+ who underwent laparoscopic intervention.

Intervention: dexmedetomidine at any posology, used either in induction, during the surgery, or after the laparoscopic device removal.

Comparison: any comparison is of interest: placebo, other anesthetics such as bupivacaine, or opioids are acceptable.

Outcome: the outcomes of interest were hospital overall death, acute renal injury (AKI) defined by the author, urinary output (ml on the first day after surgery), and LOS in days.

Study design: randomized clinical trials.

### Search strategy and information sources

The initial search strategy was performed in the bibliographic database PubMed, later adapted to Scopus, Web of Science, Cochrane trial databases, and Embase. The initial search strategy performed at PubMed was based on the health descriptors (NCBI MeSH) and the strategy was as follows: (laparoscop* OR videolaparoscop* OR "Laparoscopy"[Mesh]) AND ("Dexmedetomidine"[Mesh] OR precedex OR precedes OR dexmedetomidine) AND ("Elderly").

### Selection process and data collection process

The abstracts identified from the remote search were assessed by the two blinded authors (YNK and PEAA) between March 2023 and March 2024. The abstracts were uploaded in Rayann. These abstracts should meet at least partially the PICOS items. The disagreements were solved by consensus in weekly meetings and eventually by consulting the full manuscript. Later, the full manuscripts from all included abstracts were assessed, and their data were extracted to full forms in a blinded fashion. Disagreements were solved in consensus meetings. During the review, there was an attempt to match retrieved trial registries and full manuscripts filled out to join multiple sources from the same trial. E-mails were sent to authors in an attempt to retrieve not-available data from both trial registries and full manuscripts’ final reports.

### Data items

Data collection research forms were developed and tested in several rounds. After being ready, they were inserted into RedCap and structured with the sections: identification, eligibility, study characteristics, population characteristics, risk of bias, and outcome data.

The AKI concept of interest was initially defined as the KDIGO criteria.^[9]^ However, as this concept was rare during the assessment, any definition of AKI by the authors was accepted. Data presented in the manuscript but not in the expected format were requested from the authors. This includes LOS presented either as binary or in means and standard deviation when it was expected to be presented as medians, IQR(IQR), and range. Data was also requested when absent, for example, when in the methods section the authors mentioned that data would be analyzed but later stated to be not significant and not shown in tables or graphs; when there was clear evidence that the sample had 60+ participants but no outcome data regarding this age strata was available. These e-mails were standardized, and the research form questions were pasted in the message body to facilitate a reply with the data of interest.

### Study risk of bias assessment

Trials were evaluated with the Cochrane Risk of Bias tool version 2 (Rob2), of August 2019, and graded in “Low Risk," “High Risk,” and “Some Concerns," following the Rob2 documentation and instructions. Similar to classification steps, the two authors assessed the full manuscripts and extracted data in a blinded fashion and later solved disagreements by consensus in weekly meetings.

### Effect measures

For the outcome of overall hospital death and AKI incidence, the plan was to estimate the risk ratio; therefore, the number of participants with AKI and the total analyzed in the groups using DEX and the comparator were required. The same was performed regarding hospital mortality. For outcomes of urinary output and LOS, the plan was to estimate the standardized mean difference; therefore, when data of interest was reported as medians, it was transformed into means.

### Synthesis methods

The synthesis was planned to include all clinical trials with available data. Therefore, the clinical trials with no data would be presented with zero weight in the meta-analysis. For the AKI and death outcomes, the inverse variance method was planned to estimate the pooled summary. For the urinary output and length of stay, the standardized mean difference was used. Specifically for the LOS, in which the values distribution is known to be asymmetric, the standardized mean difference was planned to be a combination of reported means (and SDs) and means (and SDs) estimated from median and interquartile range and range using the Cai’s method for unknown non-normal distributions (MLN) approach.^[10]^ Pooled summaries were estimated with a common (fixed) effect model. Heterogeneity was assessed with the I² and Q heterogeneity test. Reporting bias was planned to be explored with a funnel plot and Egger test. All analyses were performed with *meta* and *robvis* packages in R-project software.

### Certainty assessment

The results were tabulated and evaluated using the Grading of Recommendations Assessment, Development, and Evaluation (GRADE) method and the GRADEPro web application. For each GRADE dimensions, the reviewers discussed how confident they were about the results and removed points from the dimension, later the overall confidence is assembled by considering the confidence in each dimension. GRADE structures the recommendations as strong or weak based on the confidence of the body of evidence and the balance of benefits and harms.^[11,12]^

## Results

### Study selection

The initial selection in database research returned 2,749 studies, and 652 replicates were removed, resulting in 2,097 abstracts that underwent screening by title and abstract, resulting in 45 studies included for full-text reading.

After abstract selection, 26 records were full manuscripts, and 19 were trial registries. Five studies did’t have patients aged 60+ in their sample, three studies did’t have a comparison group, and two studies did’t have any outcome of interest and were excluded. Two trial registries were replicates; only one trial registry was matched with a published manuscript. The remaining trial registries did’t have any available data on the registry website. No authors returned attempts to retrieve additional data regarding the trial registries or incomplete data from full manuscripts. In the end, there were 13 studies from which data could be extracted and analyzed (Figure 1).

**Figure 1:**
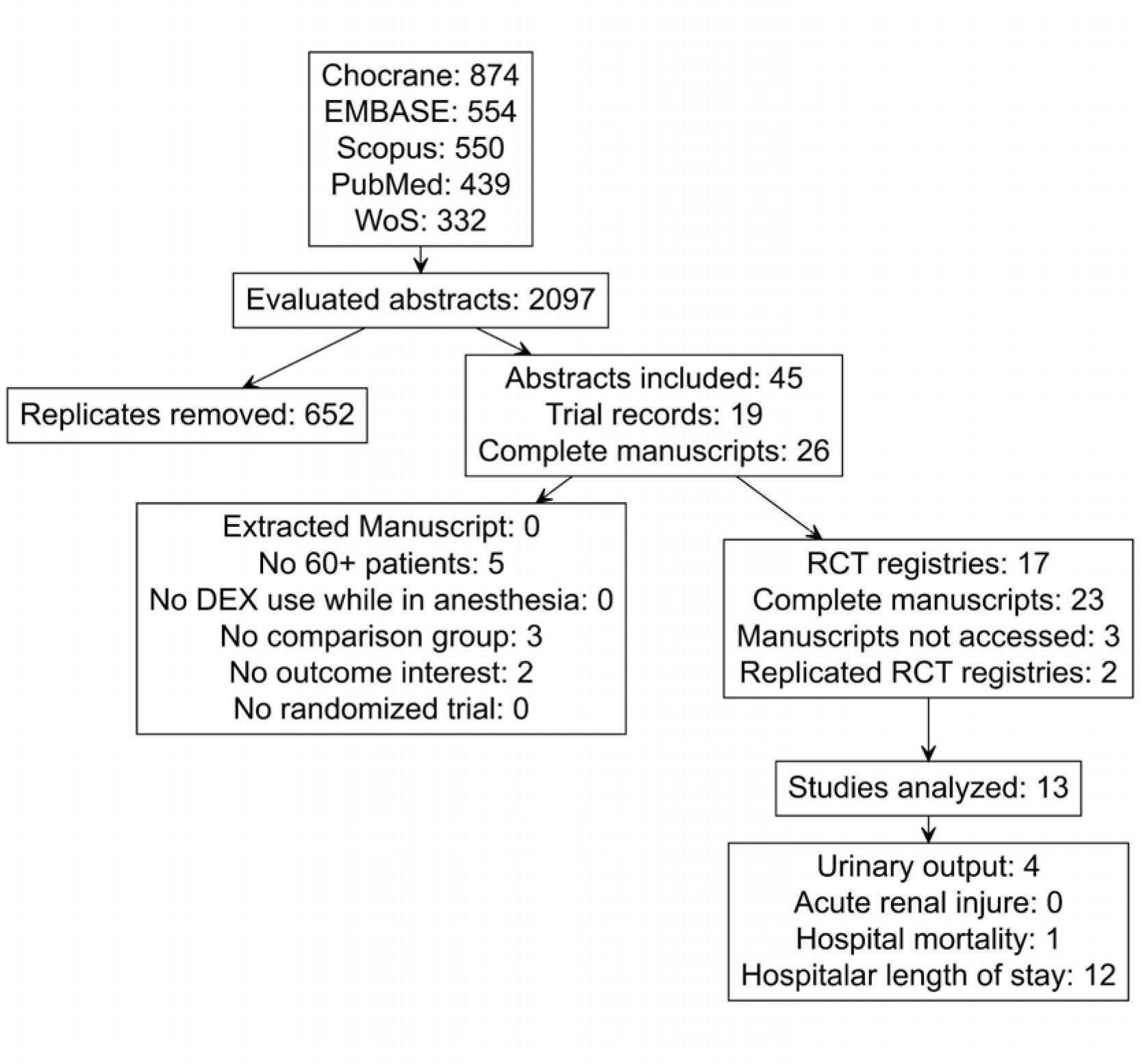
Inclusion and exclusion flow diagram

### Study characteristics

Eleven reports were from China, one from India, and one from Ukraine (Table 1). All studies were published between 2016 and 2023. Although random allocation was always mentioned, the random allocation type was’t clear in most studies, and the allocation concealment was not mentioned in almost half. Most trials mentioned double blinding, and it was always the participant and outcome measurement blinding. Many kinds of surgical clinics were involved (Table 1). The type of anesthesia varied according to the surgical proposal as follows: in eight studies, the approach was general anesthesia, in three was regional anesthesia, and one was infiltration of the portal site. The time and form of administration of DEX varied according to the anesthetic proposal and was used as an adjuvant in general anesthesia, regional anesthesia, or local anesthesia (Table 1).

**Table 1.** Descriptive results of the included studies regarding study characteristics.

| ID | Country | Enrollment period | Random allocation | Allocation concealment | Blinding | Surgery | Type of anesthesia | Anesthesia timing |
| --- | --- | --- | --- | --- | --- | --- | --- | --- |
| Chen-2016 | China | 2015-2015 | Ignored | Ignored | Double blind | Colorectal Surgery | General anesthesia | Induction / Continuous infusion |
| Bielka-2018 | Ukraine | 2016-2017 | Block random | Ignored | Single blind | General Surgery | General anesthesia | Induction / Continuous infusion |
| Selvaraj-2019 | India | - | Ignored | Sealed envelope | Double blind | General Surgery | Others | Insufflation |
| Wu-2019 | China | 2014-2015 | Ignored | Pharmacy control | Triple blind | Urological Surgery | General anesthesia | Induction / Continuous infusion |
| Pan-2020 | China | 2017-2018 | Ignored | Pharmacy control | Double blind | Colorectal Surgery | Regional anesthesia | Insufflation |
| Duan-2021 | China | 2019-2020 | Ignored | Ignored | Double blind | Oncological Surgery | General anesthesia | Induction / Continuous infusion |
| Huang-2021 | China | 2016-2017 | Ignored | Ignored | Double blind | Urological Surgery | General anesthesia | Induction / Post-surgery Continuous infusion |
| Huang-2021 | China | 2016-2017 | Ignored | Ignored | Double blind | Urological Surgery | General anesthesia | Induction / Post-surgery Continuous infusion |
| Sun-2021 | China | 2017-2018 | Ignored | Sealed envelope | Triple blind | Colorectal Surgery | General anesthesia | Continuous infusion |
| Chen-2022 | China | 2018-2022 | Ignored | Sealed envelope | Double blind | Oncological Surgery | General anesthesia | Induction / Continuous infusion |
| Jiang-2022 | China | 2014-2015 | Ignored | Sealed envelope | Triple blind | Urological Surgery | General anesthesia | Others |
| Yi-2022 | China | 2021-2022 | Ignored | Ignored | Double blind | Oncological Surgery | General anesthesia | Induction / Continuous infusion |
| Li-2023 | China | 2022-2023 | Ignored | Sealed envelope | Double blind | Urological Surgery | Regional anesthesia | Induction |

The studies usually had small sample sizes, as usually, the main outcomes were analogic scales of pain or analgesic rescue. Renal function, LOS, and hospital mortality were always secondary outcomes. The fraction of males was higher in most studies. Only one study included solely participants 60+;^[10]^ the remaining studies included a mix of participants in which there were participants at least 60 years old or younger in varying quantities. Although ASA classification was variable between the studies, patients classified as ASA II were the majority (Table 2).

**Table 2.**
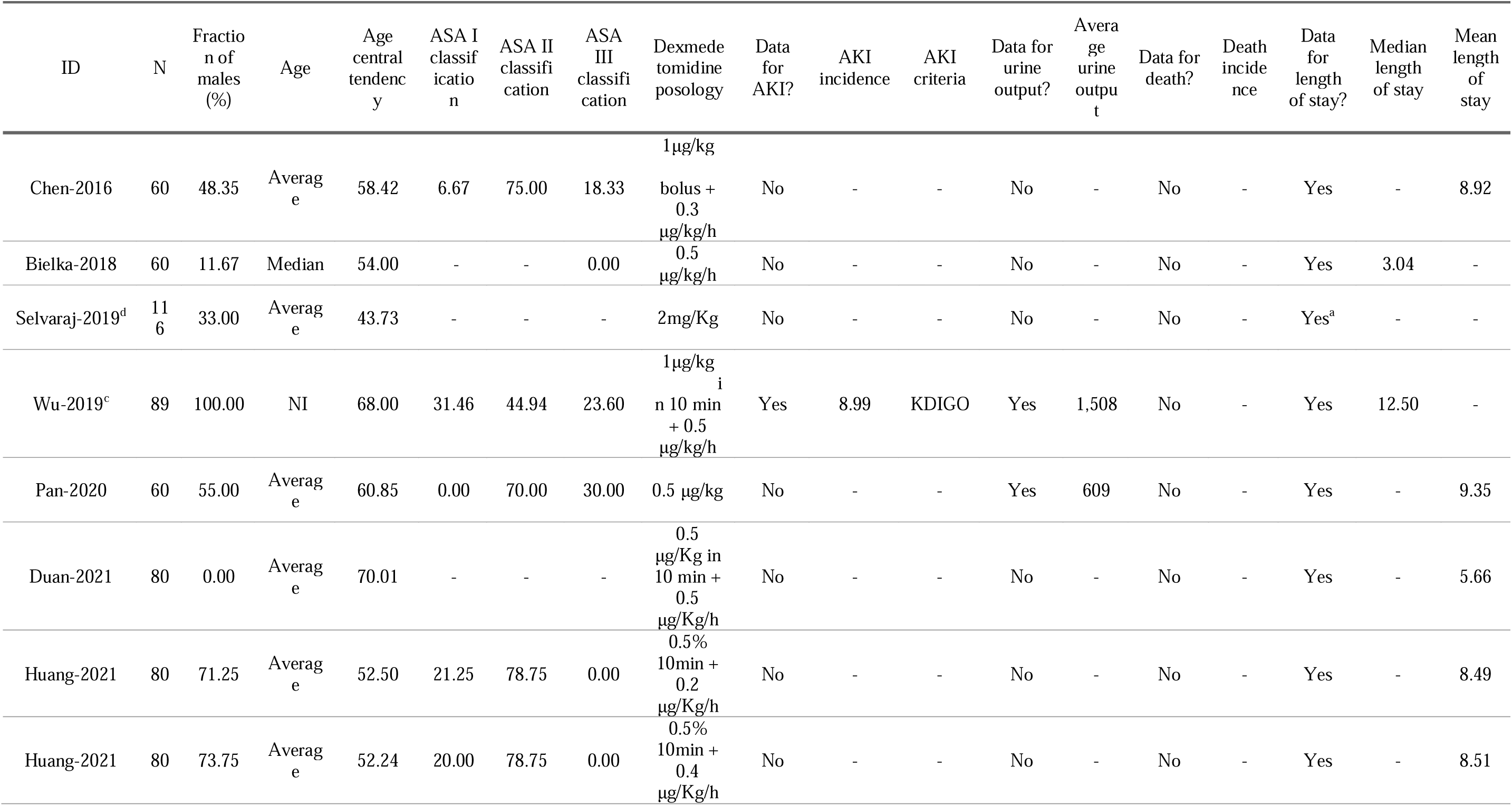
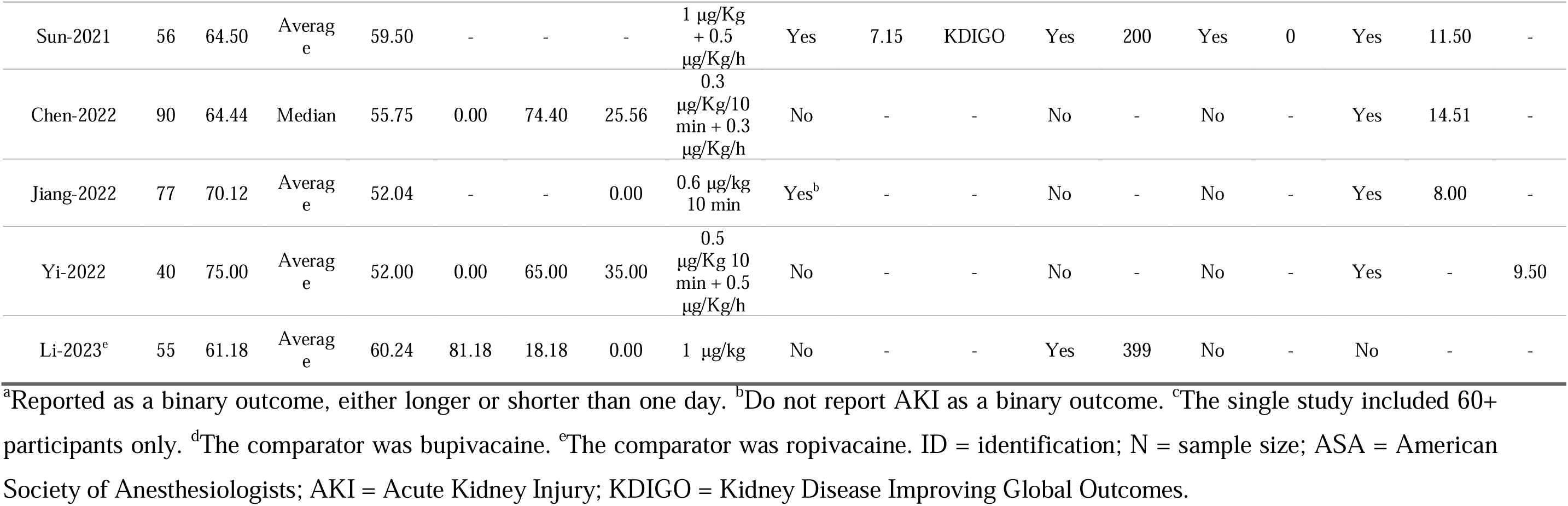
Descriptive results of the included studies regarding study sample characteristics and outcomes.

Only three studies contained information on AKI, and its overall incidence ranged from 7.15 to 8.99,^[13–15]^. Two studies defined AKI based on the diagnostic criteria established by KDIGO. Four studies provided data on urinary output on the first day after surgery (Table 2).

Only one study provided data on hospital deaths^[12]^ on the other hand, only one study did’t provide data on the overall length of hospital stay. ^[15]^ No study provided the full distribution of median and IQR. Six studies provided mean LOS and SD^[16–20]^ and five provided median LOS and its IQR.^[13,14,21–23]^ One study provided LOS data as binary (Table 2).^[15]^

Most studies used placebo as a comparator, except for one using bupivacaine as a comparator^[24]^ and one using ropivacaine as a comparator.^[22]^ The authors used many kinds of administration ways and posologies, such as intravenous continuous infusions, *in bolus,* and as adjuvant of local infiltration, and the doses varied between 0.3 μg/kg up to 1 μg/kg, which were used through single shots or as continuous infusions.

### Risk of bias in studies

The two dimensions that generated concerns of bias were the randomization process and deviations from the intended interventions. The issue regarding randomization is usually due to poorly described randomization procedures or concealment. (Figure 1: Supplementary file) The overall risk of bias assessment showed 6 studies with “Low Risk” of bias, 5 studies with “Some concerns,” and 1 study with “High Risk” of bias during the overall evaluation. (Figure 2: Supplementary file).

**Figure 2:**
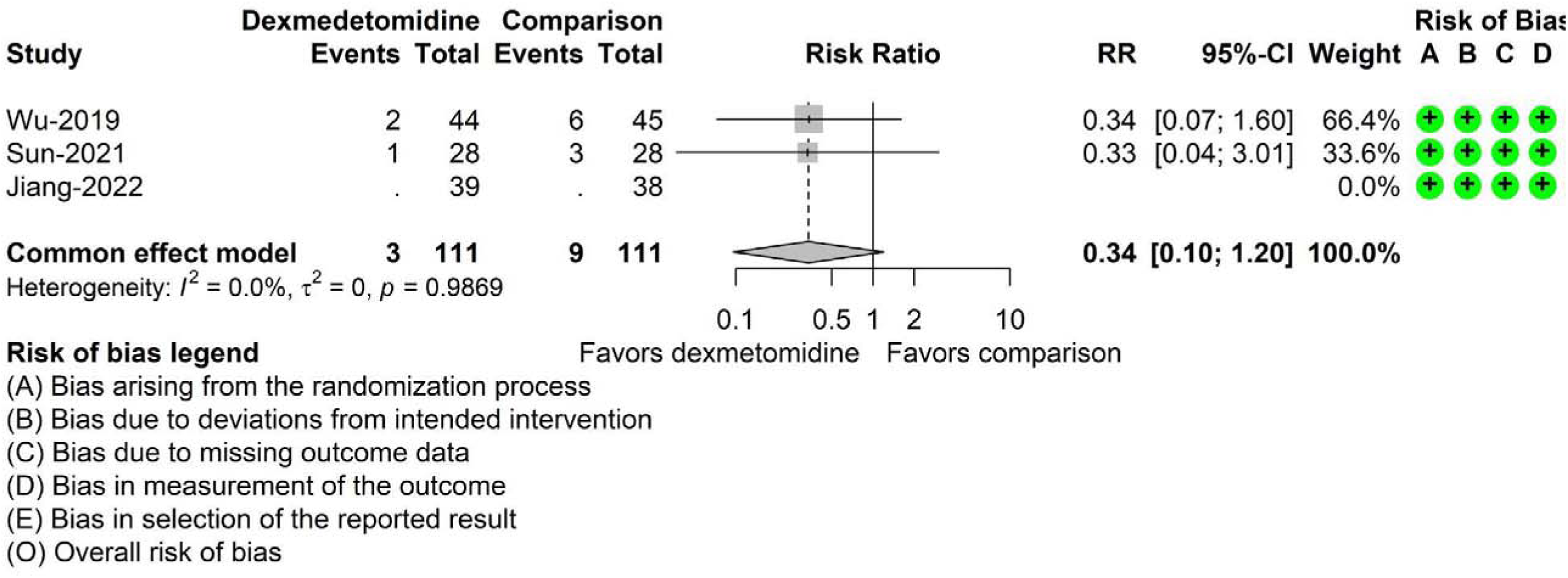
Forest plot of DEX vs. placebo effect on acute kidney injury.

### Results of individual studies

Out of the three studies reporting data regarding acute renal injury,^[14,24,25]^ only two had sufficient data to perform a meta-analysis. All three studies had an overall risk of bias assessment as low risk. Both studies with risk ratio estimates had very similar effects favoring DEX benefits in preventing AKI, consistent in the same direction and with similar precision. With this data, heterogeneity was not detected. The imprecision of the pooled estimate is large; therefore, it seems there is a DEX potential benefit in preventing AKI, but more studies designed for this purpose are required to observe benefits with reasonable precision. Reporting bias was not explored due to the small number of studies. (Figure 2)

Out of the four studies reporting data regarding urine output on the first admission day after surgery, one did’t have enough data to perform a meta-analysis^[24]^. Studies pointed standardized mean differences in different directions, showing inconsistency in data.

However, no study was able to detect a difference in urinary output among groups, and the pooled estimate also did not detect a significant effect. Heterogeneity was not detected with this data, and reporting bias was not explored due to the small number of studies. All studies included in this meta-analysis were classified as low risk of bias. (Figure 3)

**Figure 3:**
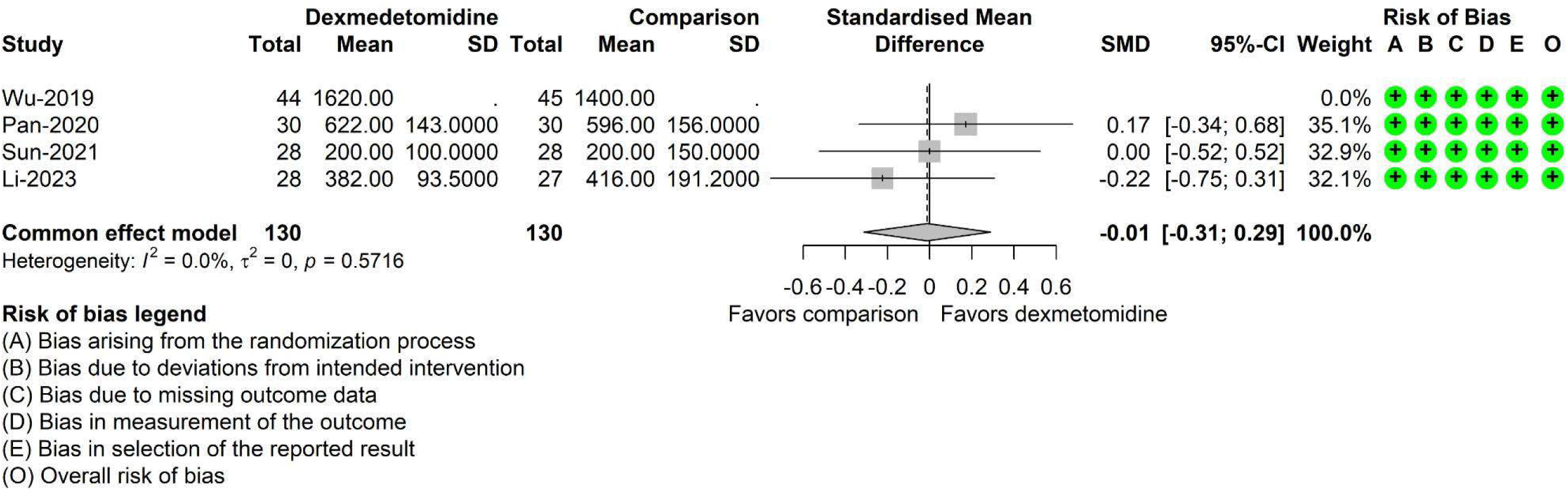
Forest plot of DEX vs. placebo effect on first-day urinary output.

Out of the twelve studies reporting data regarding hospital LOS, one did’t have sufficient data to perform meta-analisys^[26]^. Most of the studies showed a neutral effect regarding LOS. Three studies detected a benefit favoring DEX in decreasing the LOS. However, two out of these three were the smallest studies included, and two of them had some concerns regarding the risk of bias assessment, which were the one with the largest difference and the one with the largest sample size out of these three. Despite the pooled estimate showing some effect favoring the use of DEX in reducing LOS, with less than a half-day difference, the magnitude of the effect is likely to be clinically not relevant. Additionally, there is a large evidence of heterogeneity, and out of the twelve studies, five had either some concern or high risk classification in the bias assessment (Figure 4). When performing the very same analysis and selecting the seven studies with an overall classification of low risk of bias, the heterogeneity reduces about 15%, yet very high, and the pooled estimate SMD moves to a neutral effect, not favoring DEX nor the comparison (data not shown). Additionally, in this subset, the smallest study with a sample size of 54 is the most influential and the one that most contributes to heterogeneity (data not shown).

**Figure 4:**
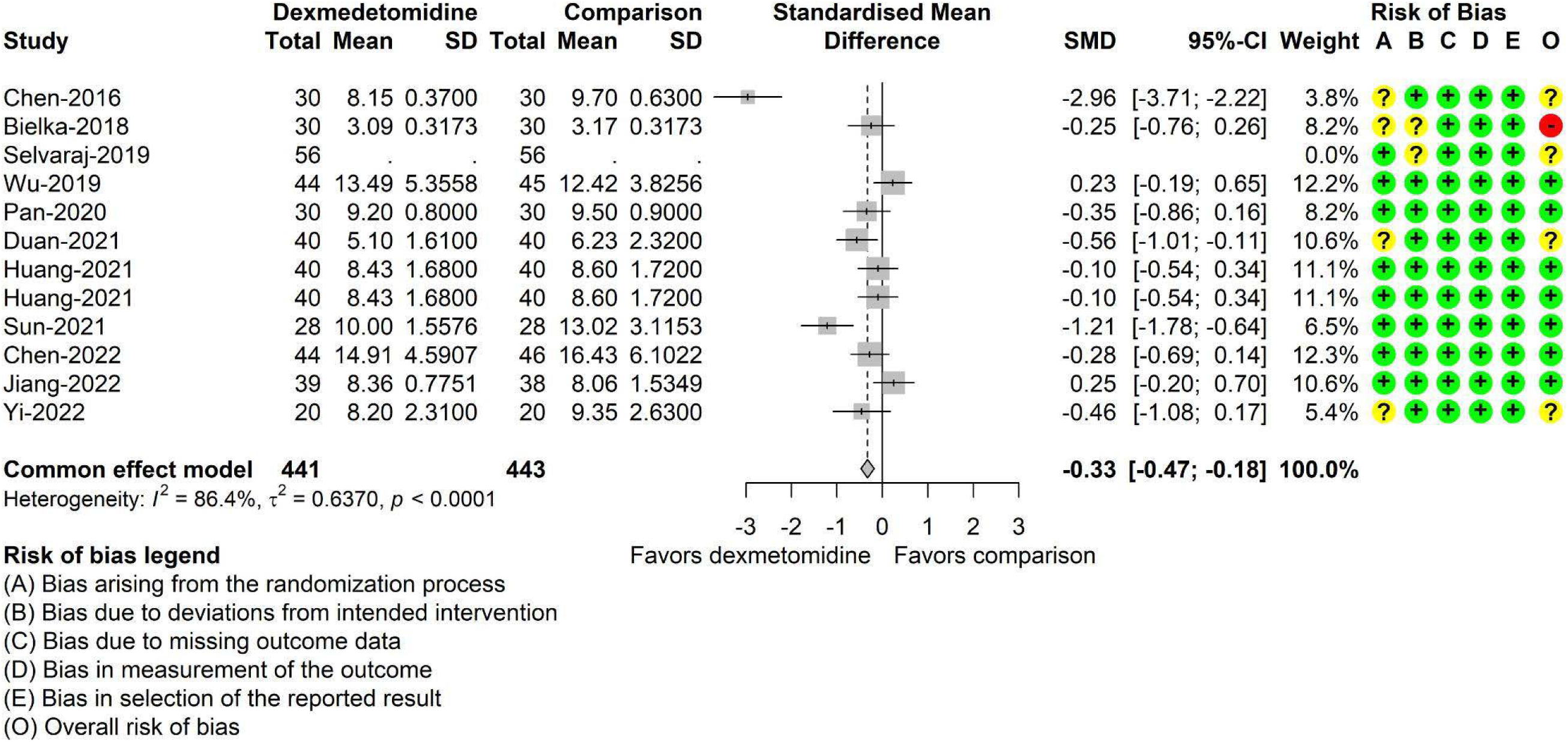
Forest plot of DEX vs. placebo effect on length of stay.

### Reporting biases

During the review process, we came across the absence of specific data for the elderly population. Only one article contained data of patients aged 60+, and all others that included this population did’t separate the data specifically for elderly patients. We attempted to contact the authors via email to obtain specific data for the population 60+, but we did’t receive any response from any of the authors whose manuscripts were included in this review.

We found a considerable number of trial registries with the potential to be included. However, none had available data on the registry website, even though many were way over the anticipated date to finish the trial.

There is also an issue regarding LOS reporting, as some manuscripts reported it as means and SD and others as median and IQR. Usually, LOS has an asymmetrical distribution and it’s skewed to the left. As the meta-analysis approach requires means and SD, in this scenario the data in the median and IQR were transformed into means and SD. However, without the range information, the means and SD estimates are more susceptible to bias.

Despite there being two studies outside the funnel at each side, there is no visually evident asymmetry in the funnel plot of DEX effect on LOS (Figure 3: Supplementary file). Nevertheless, the linear regression test of funnel plot asymmetry returned a very significant p-value of 0.0023, therefore, there are plenty of arguments and evidence for the presence of reporting bias.

### Certainty of evidence

A moderate level of confidence was found for the potential nephroprotective effect of DEX (as part of multimodal anesthesia) considering the body of evidence in studies including the elderly population undergoing laparoscopic surgery for radical prostatectomy and colorectal cancer who received DEX as part of the multimodal strategy in their anesthesia. There is plenty of evidence showing DEX has a beneficial effect on AKI incidence in other populations and other kinds of surgeries, which makes this potential benefit very plausible. The main issues reducing confidence in this finding are precision, as sample sizes were not planned to detect the benefit on AKI, and indirectness, as the effect on the elderly population is possibly mixed with the effect of the younger age group. (Table 1: Supplementary file)

The confidence of the findings regarding the effect of DEX reducing LOS is very low. The problems are on all dimensions of GRADE assessment. Additionally, even if there is a true beneficial effect of DEX in reducing LOS, the current body of evidence points to it being very unlikely to be clinically relevant, with a potential benefit of less than a third of a day than the comparison. Regarding urinary output, the confidence of findings given the current evidence is low. All GRADE dimensions for this outcome, but the risk of bias, were downgraded. Similar to LOS, the current body of evidence points to no beneficial effect of DEX on urinary output.

## Discussion

The main results to be discussed are: a) there is no evidence that intraoperative DEX in laparoscopic surgery reduces the hospital LOS in the elderly; b) there is no evidence that intraoperative DEX in laparoscopy improves urinary output on the first day of admission in the elderly; c) there is evidence of the potential nephroprotective effect of intraoperative DEX in laparoscopic surgery in the elderly. If the nephroprotector effect is true, it seems it is not related to the urinary output.

A reduction in length of hospital stay, delirium, time on mechanical ventilation, and ICU LOS was associated with DEX use in hospitalized patients, however, results appear to differ depending on the population and surgical setting. ^[27–30]^ Up to date, no studies on the elderly population have demonstrated a direct effect on reducing LOS, despite reports that it has a positive effect in the elderly population due to its anti-inflammatory action. This may be related to the pharmacological properties of the drug or possibly the timing and manner in which DEX is administered.^[27,31–33]^

Studies have shown that laparoscopy is associated with decreases in LOS, ranging from 24 to 120 hours. Notably, colorectal resections approached laparoscopically have shown more pronounced benefits in the elderly compared to younger people, including lower morbidity rates and reduced length of stay.^[34–36]^

Once statistical heterogeneity is found regarding DEX effect on LOS, one may consider clinical or methodological sources for this heterogeneity. Even after removing studies with some concerns or high-risk bias, there is still some heterogeneity. Likely, differences among populations, DEX doses, and the diversity of clinics contributed to the heterogeneity. Not at random, missing data may also lead to heterogeneity.

The quantification of urinary output involves maintaining an indwelling urinary catheter, which, according to studies, should be removed around 24-48 hours.^[37,38]^ Although there are changes in urinary patterns related to age, elderly people maintain their urinary and renal function without changes if they receive adequate volume replacement in the perioperative period.^[39]^

Studies have shown an increase in urinary output despite the use of DEX in various situations, both clinical and experimental. The mechanism behind this effect is believed to be related to blocking the release of antidiuretic hormone.^[40–42]^

The measurement of urinary output is commonly associated to renal function, it is known that in the perioperative period, several factors influence urinary output, such as the release of vasopressin and aldosterone in response to surgical stimulus. Studies have shown that urinary output is reduced in patients under general anesthesia, making urinary output an unreliable marker of renal function in this setting.^[2,43,44]^

The body of evidence in this review showed a neutral effect of DEX on urinary output. The studies analyzed show results in different directions, and the combined analysis did’t find a significant effect in any intervention.^[14,15,17]^ Although this review found that DEX may have a nephroprotective effect in the studied population, this effect is not followed by an increase in urine output. Instead, it may be assessed by novel biomarkers of acute renal failure.

Risk factors for postoperative AKI include advanced age, length of operation, and kind of surgery. AKI may also develop more quickly in the postoperative phase if there are concomitant risk factors.^[45]^ In the process of AKI, changes in serum creatinine and urea are late markers of the ongoing injury. Using urea and creatinine to diagnose AKI may delay diagnosis and appropriate treatment. The adoption of novel biomarkers of acute kidney damage, such as Cystatin-C and NGAL, may help in the early and conclusive diagnosis of AKI.^[2]^

There is evidence pointing to DEX reducing AKI and enhancing renal function in the postoperative phase of non-cardiac procedures.^[45]^ From the studies included in this review regarding AKI, it can be seen that there is no heterogeneity, and there is a non-significant but consistent potential nephroprotective effect. In this way, the use of DEX as an adjuvant in a multimodal strategy in the elderly has a potential benefit to reduce AKI and its consequences in this group of patients during the hospitalization period.

All reports included in this review didn’t match perfectly the review’s question, the population, and there are also variations regarding comparison and outcomes of interest, which often led us to encounter a lack of specific data attending to the original research question. During the analysis, we observed different dosages, routes of administration, and surgical times for the administration of DEX, which varied according to the researcher’s interest in their primary objectives. Therefore, there is a degree of indirectness in the review analysis, especially regarding population and intervention definitions. The studies that allowed the evaluation of acute renal failure and urinary output are few, six in total. They all had small sample sizes for this purpose. In addition, there is a lack of clarity regarding the definition of these outcomes as compared to the originally stated primary outcomes. It was not possible to explore reporting bias for all review outcomes, as for two of them the amount of included studies was very low for this purpose.

## Conclusion

We conclude that, based on the current data, DEX use in laparoscopies among the elderly didn’t impact the reduction of hospital LOS and shouldn’t be recommended for this purpose in this population. Similarly, DEX use in laparoscopies among the elderly didn’t improve the urinary output and shouldn’t be recommended for this purpose in this population. There is moderate confidence that DEX use in laparoscopies among the elderly has the potential to reduce AKI. To increase certainty regarding this conclusion, studies planned to use DEX as an intervention in laparoscopies among the elderly should take AKI as the primary outcome and should either include the elderly population exclusively or report results for elderly strata. AKI planned as the primary outcome will ensure that a reasonable sample size will be estimated in advance, and biomarkers to represent this outcome properly will be reasonably planned.

DEX is already regularly used in multimodal anesthesia. There is no evidence that a particular dosage in this strategy is either beneficial or may do harm in this setting for this population in preventing AKI. However, likely, similar doses to those used to reduce opioid consumption, prevent PONV, reduce shivering, and control hemodynamic parameters will be used to investigate AKI prevention.

## Data Availability

All data produced in the present work are contained in the manuscript.

## Acknowledgments

Special thanks to my colleagues at the Central Hospital from the Brazilian Army for their support during the research and to the Evandro Chagas National Institute of Infectology at the Oswaldo Cruz Foundation for the lessons learned.

## Bibliographic references

1. Hwang E, Sathianathen N, Jung J, Kim M, Dahm P, Risk M. Single-dose intravesical chemotherapy after nephroureterectomy for upper tract urothelial carcinoma. Cochrane Database of Systematic Reviews [Internet] 2019;(5). Available from: 10.1002/14651858.CD013160.pub2

2. Gumbert SD, Kork F, Jackson ML, Vanga N, Ghebremichael SJ, Wang CY, et al. Perioperative Acute Kidney Injury. Anesthesiology [Internet] 2020 [citado 2023 ago 24];132(1):180–204. Available from: https://pubs.asahq.org/anesthesiology/article/132/1/180/108774/Perioperative-Acute-Kidney-Injury

3. Kork F, Balzer F, Spies CD, Wernecke KD, Ginde AA, Jankowski J, et al. Minor Postoperative Increases of Creatinine Are Associated with Higher Mortality and Longer Hospital Length of Stay in Surgical Patients. Anesthesiology [Internet] 2015 [citado 2024 out 31];123(6):1301–11. Available from: https://pubs.asahq.org/anesthesiology/article/123/6/1301/14102/Minor-Postoperative-Increases-of-Creatinine-Are

4. Gerlach AT, Dasta JF. Dexmedetomidine: An Updated Review. Ann Pharmacother [Internet] 2007 [citado 2024 jun 19];41(2):245–54. Available from: 10.1345/aph.1H314

5. Barends CRM, Absalom A, Minnen B van, Vissink A, Visser A. Dexmedetomidine versus Midazolam in Procedural Sedation. A Systematic Review of Efficacy and Safety. PLOS ONE [Internet] 2017 [citado 2024 nov 7];12(1):e0169525. Available from: https://journals.plos.org/plosone/article?id=10.1371/journal.pone.0169525

6. Abdallah FW, Abrishami A, Brull R. The Facilitatory Effects of Intravenous Dexmedetomidine on the Duration of Spinal Anesthesia: A Systematic Review and Meta-Analysis. Anesthesia & Analgesia [Internet] 2013 [citado 2024 nov 7];117(1):271–8. Available from: https://journals.lww.com/00000539-201307000-00039

7. Duan X, Coburn M, Rossaint R, Sanders RD, Waesberghe JV, Kowark A. Efficacy of perioperative dexmedetomidine on postoperative delirium: systematic review and meta-analysis with trial sequential analysis of randomised controlled trials. British Journal of Anaesthesia [Internet] 2018 [citado 2024 nov 7];121(2):384–97. Available from: https://linkinghub.elsevier.com/retrieve/pii/S0007091218304471

8. Loomba RS, Villarreal EG, Dhargalkar J, Rausa J, Dorsey V, Farias JS, et al. The effect of dexmedetomidine on renal function after surgery: A systematic review and meta-analysis. Journal of Clinical Pharmacy and Therapeutics [Internet] 2022 [citado 2024 nov 7];47(3):287–97. Available from: https://onlinelibrary.wiley.com/doi/abs/10.1111/jcpt.13527

9. Khwaja A. KDIGO Clinical Practice Guidelines for Acute Kidney Injury. Nephron Clinical Practice [Internet] 2012 [citado 2024 jun 18];120(4):c179–84. Available from: 10.1159/000339789

10. Cai S, Zhou J, Pan J. Estimating the sample mean and standard deviation from order statistics and sample size in meta-analysis. Stat Methods Med Res 2021;30(12):2701– 19.

11. Guyatt GH, Oxman AD, Vist GE, Kunz R, Falck-Ytter Y, Alonso-Coello P, et al. GRADE: an emerging consensus on rating quality of evidence and strength of recommendations. BMJ [Internet] 2008 [citado 2024 out 22];336(7650):924–6. Available from: https://www.bmj.com/content/336/7650/924

12. Goldet G, Howick J. Understanding GRADE: an introduction. Journal of Evidence-Based Medicine [Internet] 2013 [citado 2024 out 22];6(1):50–4. Available from: https://onlinelibrary.wiley.com/doi/abs/10.1111/jebm.12018

13. Wu S, Yao H, Cheng N, Guo N, Chen J, Ge M, et al. Determining whether dexmedetomidine provides a reno-protective effect in patients receiving laparoscopic radical prostatectomy: a pilot study. Int Urol Nephrol [Internet] 2019 [citado 2023 ago 24];51(9):1553–61. Available from: http://link.springer.com/10.1007/s11255-019-02171-9

14. Sun W, Li F, Wang X, Liu H, Mo H, Pan D, et al. Effects of Dexmedetomidine on Patients Undergoing Laparoscopic Surgery for Colorectal Cancer. J Surg Res 2021;267:687–94.

15. Li Y, Zhang L, Jiao J, Yu X, Huang S. Impact of Bilateral Quadratus Lumborum Block Using Different Doses of Dexmedetomidine for Postoperative Analgesia in Laparoscopic Myomectomy: A Randomized Controlled Trial. Clin J Pain 2023;39(2):85– 90.

16. Chen C, Huang P, Lai L, Luo C, Ge M, Hei Z, et al. Dexmedetomidine improves gastrointestinal motility after laparoscopic resection of colorectal cancer: A randomized clinical trial. Medicine (Baltimore) 2016;95(29):e4295.

17. Pan W, Liu G, Li T, Sun Q, Jiang M, Liu G, et al. Dexmedetomidine combined with ropivacaine in ultrasound-guided tranversus abdominis plane block improves postoperative analgesia and recovery following laparoscopic colectomy. EXPERIMENTAL AND THERAPEUTIC MEDICINE 2020;19(4):2535–42.

18. Duan JQ, Huang HG, Shui CL, Jiang W, Luo X. The cardioprotective effect of dexmedetomidine on elderly patients with cervical cancer in the trendelenburg position. Signa Vitae [Internet] 2021;17(5):117–21. Available from: https://www.embase.com/search/results?subaction=viewrecord&id=L2013825184&from=export

19. Huang SS, Song FX, Yang SZ, Hu S, Zhao LY, Wang SQ, et al. Impact of intravenous dexmedetomidine on postoperative bowel movement recovery after laparoscopic nephrectomy: A consort-prospective, randomized, controlled trial. World J Clin Cases 2021;9(26):7762–71.

20. Ou Y, Liu G, Yin F, Yang Y, Zhang F. [Protective effect of ulinastatin combined with dexmedetomidine against hepatic ischemia-reperfusion injury in laparoscopic hepatectomy for liver cancer and cirrhosis: a randomized controlled trial]. Nan Fang Yi Ke Da Xue Xue Bao 2022;42(12):1832–8.

21. Bielka, Kateryna, Kuchyn, Iurii, Babych, Volodymyr, Martycshenko, Kseniia, Inozemtsev, Oleksii. Dexmedetomidine Infusion as an Analgesic Adjuvant During Laparoscopic Cholecystectomy. https://clinicaltrials.gov/show/NCT03211871 [Internet] 2018;18(44):1–6. Available from: https://www.cochranelibrary.com/central/doi/10.1002/central/CN-01495523/full

22. Chen YX, Du L, Wang LN, Shi YY, Liao M, Zhong M, et al. Effects of Dexmedetomidine on Systemic Inflammation and Postoperative Complications in Laparoscopic Pancreaticoduodenectomy: A Double-blind Randomized Controlled Trial. World J Surg 2023;47(2):500–9.

23. Jiang Z, Zhou G, Song Q, Bao C, Wang H, Chen Z. Effect of Intravenous Oxycodone in Combination With Different Doses of Dexmedetomidine on Sleep Quality and Visceral Pain in Patients After Abdominal Surgery: A Randomized Study. CLINICAL JOURNAL OF PAIN 2018;34(12):1126–32.

24. Wu S, Yao H, Cheng N, Guo N, Chen J, Ge M, et al. Determining whether dexmedetomidine provides a reno-protective effect in patients receiving laparoscopic radical prostatectomy: a pilot study. Int Urol Nephrol 2019;51(9):1553–61.

25. Jiang L, Zhang T, Zhang Y, Yu D, Zhang Y. Dexmedetomidine postconditioning provides renal protection in patients undergoing laparoscopic partial nephrectomy: A randomized controlled trial. Front Pharmacol 2022;13:988254.

26. Selvaraj V, Kamaraj R. Effect of dexmedetomidine as an adjuvant to 0.25% bupivacaine for local infiltration of port site in laparoscopic cholecystectomy in terms of quality and duration of post-op analgesia. Anestezi Derg [Internet] 2019;27(3):210–6. Available from: https://www.embase.com/search/results?subaction=viewrecord&id=L2002540099&from=export

27. Lin C, Tu H, Jie Z, Zhou X, Li C. Effect of Dexmedetomidine on Delirium in Elderly Surgical Patients: A Meta-analysis of Randomized Controlled Trials. Ann Pharmacother [Internet] 2021 [citado 2024 nov 30];55(5):624–36. Available from: https://journals.sagepub.com/doi/10.1177/1060028020951954

28. Patanwala AE, Erstad BL. Comparison of Dexmedetomidine Versus Propofol on Hospital Costs and Length of Stay. J Intensive Care Med [Internet] 2016 [citado 2024 nov 30];31(7):466–70. Available from: https://journals.sagepub.com/doi/10.1177/0885066614544452

29. Heybati K, Zhou F, Ali S, Deng J, Mohananey D, Villablanca P, et al. Outcomes of dexmedetomidine versus propofol sedation in critically ill adults requiring mechanical ventilation: a systematic review and meta-analysis of randomised controlled trials. British Journal of Anaesthesia [Internet] 2022 [citado 2024 nov 30];129(4):515–26. Available from: https://linkinghub.elsevier.com/retrieve/pii/S000709122200321X

30. Peng K, Ji F hai, Liu H yue, Zhang J, Chen Q cai, Jiang Y hui. Effects of Perioperative Dexmedetomidine on Postoperative Mortality and Morbidity: A Systematic Review and Meta-analysis. Clinical Therapeutics [Internet] 2019 [citado 2024 nov 30];41(1):138-154.e4. Available from: https://linkinghub.elsevier.com/retrieve/pii/S0149291818305496

31. Kim DJ, Kim SH, So KY, Jung KT. Effects of dexmedetomidine on smooth emergence from anaesthesia in elderly patients undergoing orthopaedic surgery. BMC Anesthesiol [Internet] 2015 [citado 2024 nov 30];15(1):139. Available from: http://bmcanesthesiol.biomedcentral.com/articles/10.1186/s12871-015-0127-4

32. Zeng H, Li Z, He J, Fu W. Dexmedetomidine for the prevention of postoperative delirium in elderly patients undergoing noncardiac surgery: A meta-analysis of randomized controlled trials. PLoS ONE [Internet] 2019 [citado 2024 nov 30];14(8):e0218088. Available from: https://dx.plos.org/10.1371/journal.pone.0218088

33. Lee C, Lee CH, Lee G, Lee M, Hwang J. The effect of the timing and dose of dexmedetomidine on postoperative delirium in elderly patients after laparoscopic major non-cardiac surgery: A double blind randomized controlled study. J Clin Anesth [Internet] 2018;47:27–32. Available from: 10.1016/j.jclinane.2018.03.007

34. Loureiro ER, Klein SC, Pavan CC, Almeida LDLF, Silva FHPD, Paulo DNS. Colecistectomia videolaparoscópica em 960 pacientes idosos. Rev Col Bras Cir [Internet] 2011 [citado 2024 nov 30];38(3):155–60. Available from: http://www.scielo.br/scielo.php?script=sci_arttext&pid=S0100-69912011000300003&lng=pt&tlng=pt

35. Bàllesta López C, Cid JA, Poves I, Bettónica C, Villegas L, Memon MA. Laparoscopic surgery in the elderly patient. Surg Endosc [Internet] 2003 [citado 2024 nov 30];17(2):333–7. Available from: https://link.springer.com/10.1007/s00464-002-9056-7

36. Frasson M, Braga M, Vignali A, Zuliani W, Di Carlo V. Benefits of Laparoscopic Colorectal Resection Are More Pronounced in Elderly Patients. Diseases of the Colon & Rectum [Internet] 2008 [citado 2024 nov 30];51(3):296–300. Available from: https://journals.lww.com/00003453-200851030-00005

37. Nollen J, Pijnappel L, Schoones JW, Peul WC, Van Furth WR, BrunsveldlJReinders AH. Impact of early postoperative indwelling urinary catheter removal: A systematic review. Journal of Clinical Nursing [Internet] 2023 [citado 2024 nov 30];32(9–10):2155–77. Available from: https://onlinelibrary.wiley.com/doi/10.1111/jocn.16393

38. Zhang W, Liu A, Hu D, Xue D, Li C, Zhang K, et al. Indwelling versus Intermittent Urinary Catheterization following Total Joint Arthroplasty: A Systematic Review and Meta-Analysis. PLoS ONE [Internet] 2015 [citado 2024 nov 30];10(7):e0130636. Available from: https://dx.plos.org/10.1371/journal.pone.0130636

39. Kumle B, Boldt J, Suttner S, Piper SN. [Change in kidney function in elderly patients in the perioperative phase]. Med Klin (Munich) 2001;96(4):202–7.

40. Villela NR, Nascimento Júnior P do, Carvalho LR de, Teixeira A. Efeitos da dexmedetomidina sobre o sistema renal e sobre a concentração plasmática do hormônio antidiurético: estudo experimental em cães. Rev Bras Anestesiol [Internet] 2005 [citado 2024 nov 30];55:429–40. Available from: https://www.scielo.br/j/rba/a/WtPtrd9bLbsjCVr7Z74fHvP/?lang=pt

41. Granger S, Ninan D. Intraoperative Dexmedetomidine-Induced Polyuric Syndrome. Cureus [Internet] 2017 [citado 2024 nov 30];Available from: http://www.cureus.com/articles/6969-intraoperative-dexmedetomidine-induced-polyuric-syndrome

42. Fialho L, Cunha-E-Silva JA, Santa-Maria AF, Madureira FA, Iglesias AC. Comparative study of systemic early postoperative inflammatory response among elderly and non-elderly patients undergoing laparoscopic cholecystectomy. Rev Col Bras Cir [Internet] 2018 [citado 2024 nov 30];45(1). Available from: http://www.scielo.br/scielo.php?script=sci_arttext&pid=S0100-69912018000100161&lng=en&tlng=en

43. Hahn RG. Volume kinetics for infusion fluids. Anesthesiology 2010;113(2):470–81.

44. Matot I. Effect of the Volume of Fluids Administered on Intraoperative Oliguria in Laparoscopic Bariatric Surgery: A Randomized Controlled Trial. Arch Surg [Internet] 2012 [citado 2024 nov 30];147(3):228. Available from: http://archsurg.jamanetwork.com/article.aspx?doi=10.1001/archsurg.2011.308

45. Zhuang K, Yang H tian, Long Y qin, Liu H, Ji F hai, Peng K. Dexmedetomidine and acute kidney injury after non-cardiac surgery: A meta-analysis with trial sequential analysis. Anaesthesia Critical Care & Pain Medicine [Internet] 2024 [citado 2024 dez 2];43(3):101359. Available from: https://www.sciencedirect.com/science/article/pii/S2352556824000171

