## Supplemental figures for "Effects of dexmedetomidine use on elderly patient’s renal function submitted to laparoscopic surgery: systematic review"

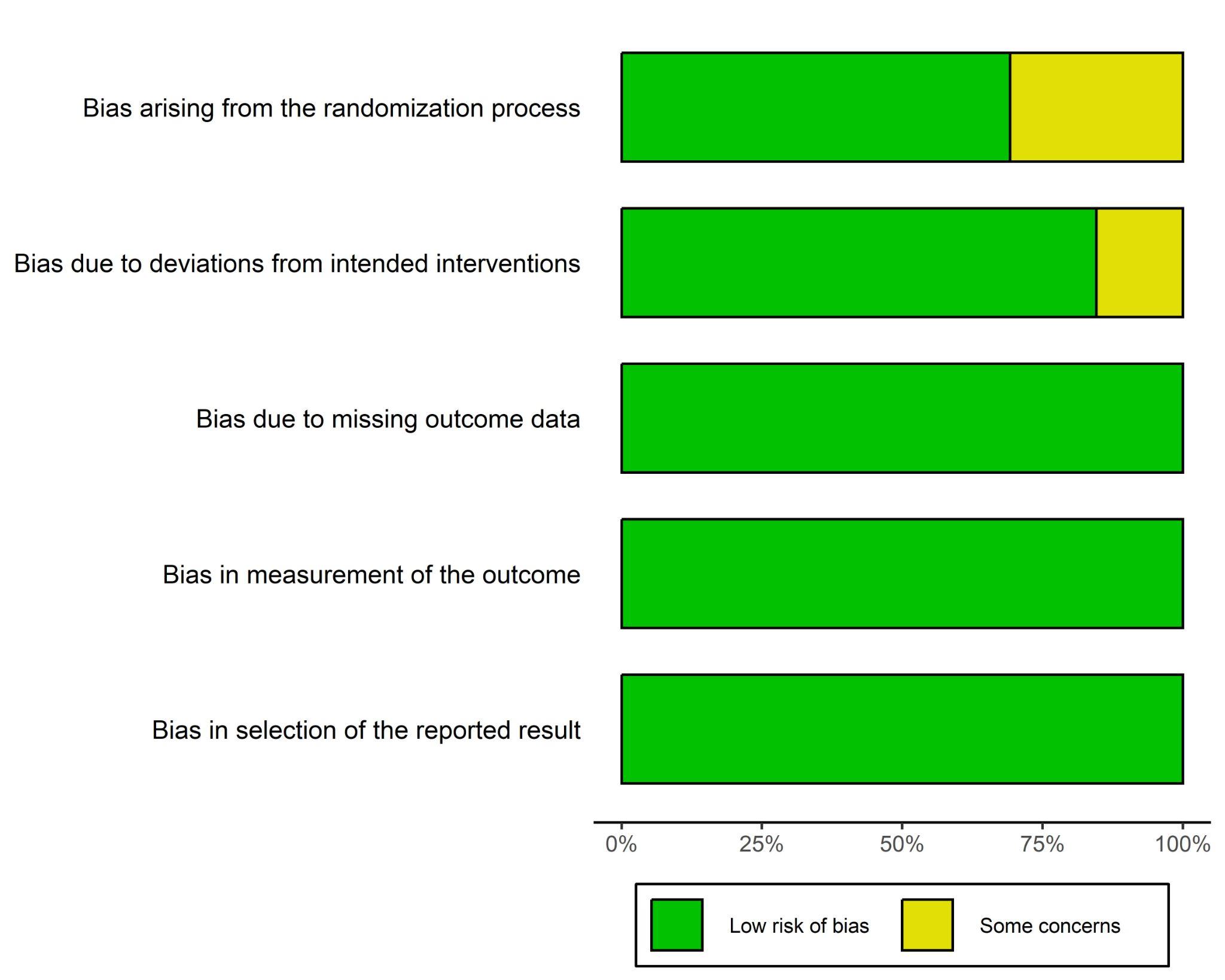

### **Figure 1:** Risk of bias with RoB2 assessment showing the proportion of studies classified within each category.

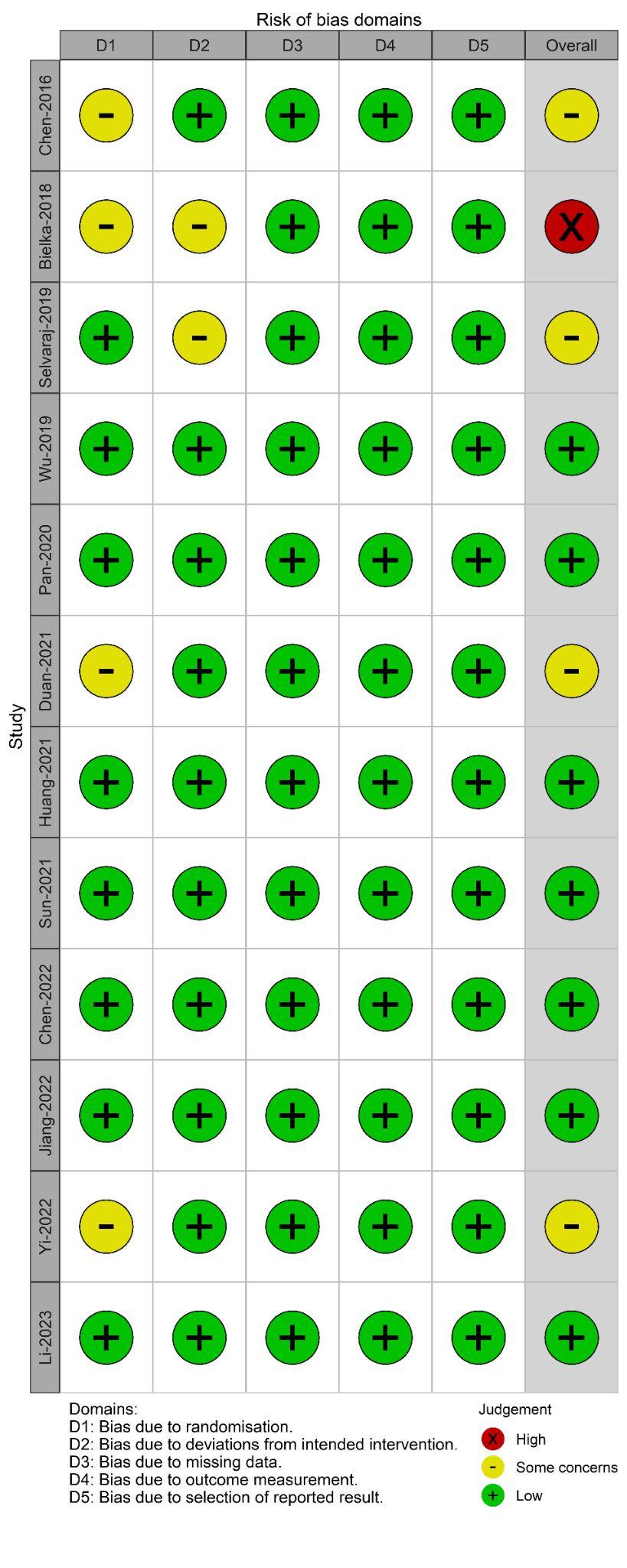

### **Figure 2**: Risk of bias with RoB2 assessment showing individual study assessment in each dimension and overall.

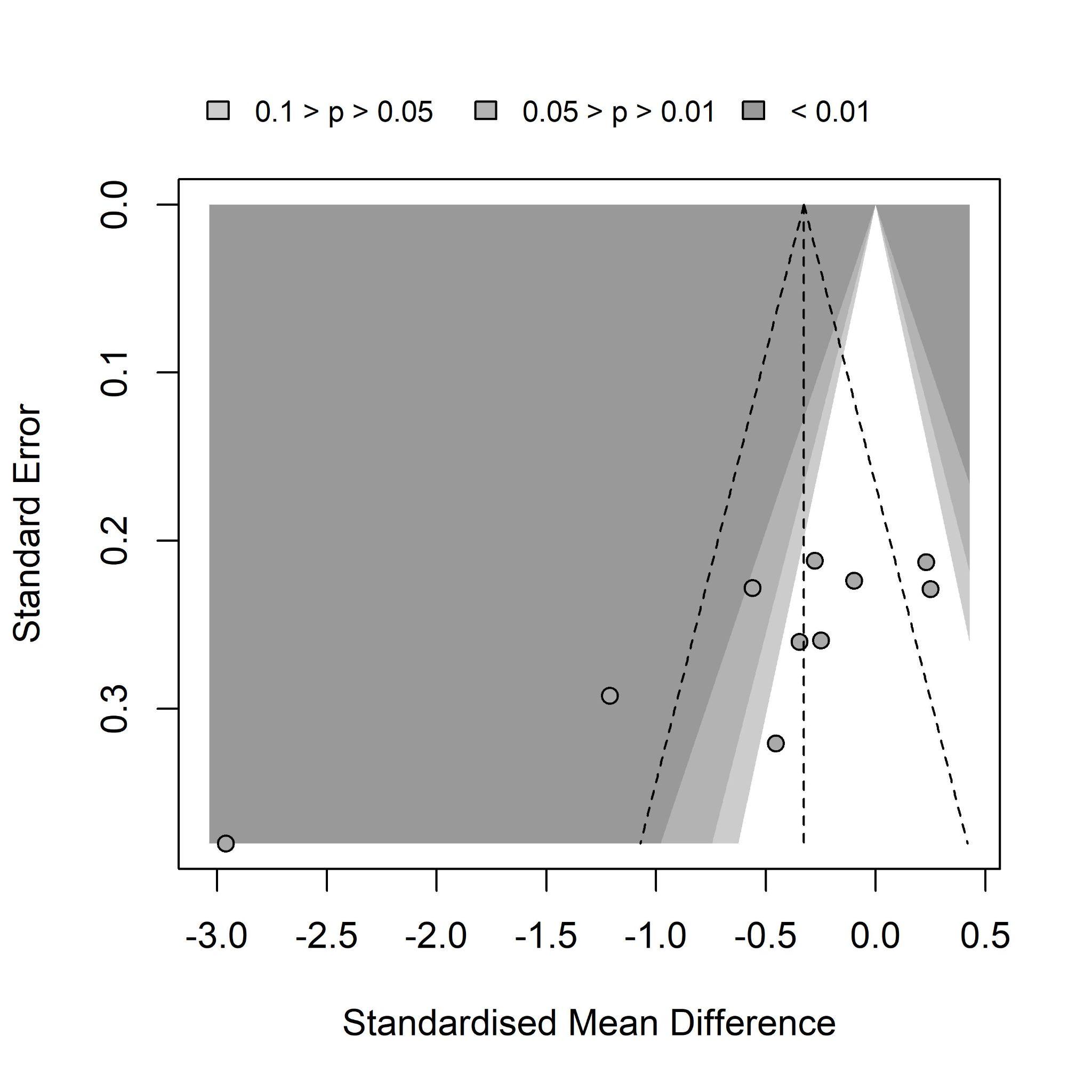

**Figure 3:** Funnel plot of DEX vs placebo effect on the LOS outcome.

**Table 1**: Summary of findings for Dexmedetomidine vs. placebo to reduce acute renal injury in elderly patients submitted

to laparoscopic surgeries.

| Outcome and follow-up | Patients  (studies), N | Relative effect  (95% CI) | Absolute effects (95% CI) | | | Certainty | What happens |
| --- | --- | --- | --- | --- | --- | --- | --- |
|  |  |  | Placebo | Dexmedetomidine | Difference |  |  |
| Acute renal injury  Follow-up: variation  3.04 days to 14.01  days | 222  (2 RCTs) | **RR = 0.34**  (0.10 to 1.20) | 81 per 1.000 | **28 per 1.000**  (8 to 97) | **54 less per 1,000**  (from 73 less to 16 more) | ⨁⨁⨁◯  Moderate (a,b) | Dexmedetomidine (DEX) during laparoscopic (radical prostatectomy or colorectal cancer) in patients between 40 and 79 years old, with ASA classification either I, II, or III, and no previous kidney, lung, liver, or heart condition, with no antitumor therapy, the use of 1 μg/kg  of DEX was intravenously administered within 10 min, followed by 0.5 μg/kg/h of DEX infusion during the laparoscopic operation, which provides a potential nephroprotective effect. |
| Length of hospital stay  Follow-up: variation  3.04 days to 14.01  days | 884  (11 RCTs) | - | - | - | **-0.32**  (-0.47 to -0.18) | ⨁◯◯◯  Very  low (c,d,e,f,g) | Dexmedetomidine (DEX) during laparoscopic (cholecystectomy, nephrectomy, pancreaticoduodenectomy, colorectal cancer) in patients between 18 and 79 years old, with ASA from I to III, and no previous kidney, lung, liver, or heart condition, with no antitumor therapy nor diabetes, the use of 0.6 μg/kg/h of IV DEX bolus or 0.3 μg/kg/h of IV DEX infusion or 1 μg/kg of IV DEX within 10 min plus 0.5 μg/kg/h of IV DEX infusion is unlikely to have any clinically relevant effect on LOS. |
| Urine output  Follow-up: median 1 day | 260  (3 RCTs) | - | - | **-** | **-0.01**  (-0.31 to 0.29) | ⨁◯◯◯  Very  low  (h,i,j,k) | Dexmedetomidine (DEX) during laparoscopic (colectomy, nephrectomy, and colorectal cancer) in patients between 18 and 75 years old, with ASA from I to II, and no previous kidney, lung, liver, or heart condition, with no antitumor therapy and no diabetes, the use of 0.5 μg/kg/h of DEX TAP block bolus or 1 μg/kg/h of DEX quadratus lumborum block bolus or 1 μg/kg of IV DEX within 10 min plus 0.5 μg/kg/h of IV DEX infusion has no clinically relevant effect on first-day urinary output. |

CI: Confidence interval; RR: Risk ratio; SMD: Standardized mean difference

a. Sun-2021, different from the other included trials, has about half of the sample with participants who are 60 years old or younger. Different dexmedetomidine posologies and routes of administration vary among trials. Participants from different surgery clinics may have different baseline prognoses.

b. All included trials state AKI as a secondary outcome. The sample size was not originally estimated to detect the effect on AKI. Additionally, there were very few reports regarding the AKI outcome, and one of them did not inform it as a binary outcome. To detect such Risk Ratio with such a small number of events among the controls would require at least 1200 overall participants, depending on the desired precision and significance.

c. For the LOS outcome, four out of the 11 studies had at least some concern about the risk of bias assessment. One of the studies had a concerning overall risk of bias assessment, although it had a neutral effect. The sensibility analysis, including only the low risk of bias studies, changed the interpretation from beneficial to no efficacy in reducing LOS.

d. For the LOS outcome, there is moderate evidence of heterogeneity, and there are trials with effects in different directions, although most of them are not significant in either direction. The significant trials favor DEX only.

e. For the LOS outcome, all included trials had mixed samples of 60+ and 60- participants. Additionally, different posologies and routes of administration are present, and every trial had patients from different surgery clinics with different baseline prognoses, which might influence the recovery and LOS in different ways.

f. LOS outcome was always a secondary outcome, and sample sizes were not planned to detect LOS differences. Eight out of the eleven trials included the null effect into their confidence interval. The estimated sample size with a 0.32 SMD varied from 880 to many thousands in both groups, depending on power, alpha, and standard deviation simulated values.

g. Publication bias was statistically detected for the LOS outcome, and there are many arguments for that. Most of the trials had partial data of interest; 1 was removed from the meta-analysis, and many of them had their means and standard deviation estimated from medians and IQR, which presumably more prompt to estimation bias when compared to when total range is also available.

h. Each of the three included trials showed different effect directions. Additionally, the single trial that had data regarding both acute renal injury and urinary output showed reduced AKI but no differences regarding urinary output among groups.

i. Different dexmedetomidine posologies and routes of administration vary among trials. Additionally, trials were performed with different surgery clinics, likely to have different control of the urinary output.

j. The observed difference in urinary output is so small that it is unlikely the presence of any clinically relevant effect. With current evidence, if the effect is real, it is marginal, and it would require many thousands of overall participants to detect such an effect.

k. Although reporting bias was not statistically detected, Wu-2019 was the only trial with 60+ participants exclusively, but it was incomplete regarding urinary output
